# Establishing consensus on emergency department interventions that could be conducted in sub-acute care settings for non-emergent paramedic transported visits: A RAND/UCLA modified Delphi study

**DOI:** 10.1101/2021.06.01.21258191

**Authors:** Ryan P Strum, Walter Tavares, Andrew Worster, Lauren E Griffith, Andrew P Costa

**Author notes:** **Corresponding Author**: Ryan P Strum, McMaster University, CRL B106, 1280 Main Street West, Hamilton, ON, L8S 4L8. **Funding Statement**: The investigators received no specific funding for this study. **Declaration of Competing Interests**: All authors of this manuscript do not declare any competing interests, financial or otherwise.

## Abstract

**Background:** Patients transported by paramedics for non-emergent conditions are increasing in Ontario and contribute to an emergency department (ED) crisis. Redirecting certain patients to sub-acute healthcare may be beneficial and suitable. We examined if ED interventions conducted on non-emergent paramedic transported patients could be conducted in sub-acute health centres.

**Methods:** A RAND/UCLA modified Delphi study was conducted. Twenty emergency and primary care physicians rated the suitability of the 150 most frequently recorded interventions for completion in sub-acute healthcare centres and provided comments to augment ratings. Interventions were performed on non-emergent adult patients transported by paramedics to an ED, and abstracted from the National Ambulatory Care Reporting System database (January 1, 2014 to March 31, 2018). We used two rounds of a modified Delphi process and set consensus at 70% agreement.

**Results:** Consensus was reached on 146 (97.3%) interventions; 103 interventions (68.7%) were suitable for sub-acute centres, 43 (28.7%) for ED only; 4 (2.6%) did not receive consensus. For sub-acute centres, all 103 interventions were rated for urgent care centres; walk-in medical centres were applicable for 46 (30.6%) and nurse practitioner-led clinics for 47 (31.3). Diagnostic imaging availability, physician preferences and staffing were determining factors for discrepancies in sub-acute centre ratings.

**Interpretation:** The majority of included ED interventions performed on non-emergent patients transported by paramedics were identified as suitable for urgent care clinics, with one-third being suitable for either walk-in medical centres or nurse practitioner-led clinics. In combination with additional patient details and supports, knowledge of interventions suitable for sub-acute healthcare centres will inform a patient classification model for paramedic-initiated redirection of patients from ED.

**Study registration:** ID ISRCTN22901977.

## BACKGROUND

Paramedic services are increasingly transporting patients with non-emergent conditions to the emergency department (ED) when primary healthcare facilities may be more beneficial for their care.(1,2) In Ontario, patients with non-emergent conditions account for 60% of all ambulance transported patients, of which 74% are discharged the same day.(3) Initiatives in health policy and research have not decreased paramedic transports for non-emergent ED visits; from 2014 to 2017 usage has increased by 12% (456 510 / 511 801) in Ontario.(4) Increasing ED visits have outpaced population growth in Ontario by more than double (13.6%, vs. 6.2% respectively)(5), suggesting utilization of ED’s has broadened. Broadened use of paramedic services by non-emergent patients and a legislative requirement to transport all patients to the ED regardless of acuity is exacerbating the problem.(6,7)

Redirecting non-emergent patients to sub-acute care centres instead of EDs may offer a feasible solution to prevent some non-emergent patient visits.(8) Patient redirection has been successful in Canada; a computer algorithm to direct non-emergent visits from ED to primary care centres not only left patients as satisfied with the care they received (84%), but was also described as a safe strategy (5.9% of 980 diverted patients had unexpected healthcare visits to the ED; none for severe complication).(9) Internationally, sub-acute centres such as urgent care clinics have reduced the likelihood of ED visits for lower acuity conditions, have shown that they can perform treatments equivalent to EDs for minor illnesses and traumatic injuries, and at a lower cost.(10–13) Redirection to sub-acute care centres by paramedics may have beneficial long-term implications by reducing paramedic transport consumption and can have higher cost effectiveness than transport to an acute centre ED.(14–17)

Evidence to support redirecting patients transported by paramedics to sub-acute centres is inconclusive, and international findings may not be generalizable across Canada.(18) Part of the difficulty arises from an absence of a suitable patient classification for examining which patients transported by paramedic services could have been potentially redirected.(17) Therefore, the objective of this study was to establish consensus on a set of ED interventions performed on non-emergent patients transported by paramedics that could be conducted in sub-acute healthcare centres.

## METHODS

We used a three-stage RAND/UCLA modified Delphi study design to evaluate consensus on ED physician interventions that could be conducted in alternative sub-acute health centres. This methodology allowed us to assess a collective groups judgements on patient procedures and facilitate group discussion between rounds.(19) The CHERRIES reporting guideline was followed to report this study.(20)

### Stage 1: Identifying ED inventions to include in the RAND/UCLA modified Delphi

We generated a list of unique Canadian Classification for Health Interventions (CCI) *main intervention* codes recorded on non-emergent adult patients (18 years or greater) that were transported to hospital by paramedics in Ontario from January 1, 2014 to March 31, 2018 from the National Ambulatory Care Reporting System (NACRS) ED database. NACRS contains an Ontario population-level collection of hospital administrative records. Non-emergent patients were considered to have a Canadian Triage and Acuity Score (CTAS) of three (urgent) to five (non-urgent), and were assigned their score upon entry to the ED by an ED or triage nurse. CTAS is an ordinal scale that ranges from one to five, with a score of one representing the most emergent (resuscitation) and five as least urgent (non-urgent). We determined *a priori* that our intervention list for the modified Delphi exercise should encompass at least 95% of total interventions in the study cohort to increase face validity.

### Stage 2: Selection of experts to constitute the Delphi expert committee

We used purposive sampling to select 25 primary care and emergency physicians who were currently or recently practicing in Ontario, Canada.(3) We sought physicians who had either extensive medical experience, academic experience, or a leadership role in paramedic practice oversight to ensure they could offer high quality comprehension when evaluating ED interventions. All selected experts were sent a study information package (objective, purpose, contribution), and those who participated gave informed consent prior to beginning the modified Delphi. We only included physicians as all interventions were performed by a physician. All other types of practitioners (including paramedics) were excluded to reduce any potential bias of experts evaluating interventions that may not be within the practitioner’s scope of practice. We determined *a priori* the Delphi expert committee must be composed of at least ten physicians, with representation from both emergency and primary care disciplines to increase the reliability of group judgements.(21) Once an expert was recruited, they were asked to complete a demographic questionnaire. Only physicians who completed at least one round would be included in the Delphi expert committee, and were provided a $75 e-gift card for participation.

### Stage 3: Exploring consensus on ED interventions that could be conducted in sub-acute health care centres

We used the RAND/UCLA modified Delphi technique to assess expert consensus through two rounds of ratings over an eight-week timeframe.(3) The modified Delphi method is a strategy that analyzes collective expert judgements to produce superior results than any one expert would, resulting in increased content validity.(22) We used a secure and encrypted *CheckMarket* software program to develop and administer the study questionnaires to experts. All data were stored on the investigators in encrypted servers. Round 1 of the modified Delphi included all ED interventions identified in Stage 1. Interventions were presented in six subsections based on their section of the CCI Tabular List, 2018 Volume 3 categorization: (1) physical/physiological therapeutic interventions; (2) diagnostic interventions; (3) diagnostic imaging interventions, and; (6) cognitive, psychosocial and sensory therapeutic interventions, (7) other healthcare interventions, and (8) therapeutic interventions strengthening the immune system.(23) Section (5) obstetrical and fetal interventions were not included as no interventions assigned in this section was identified in the cohort, and no randomization of questions were used. For each intervention, experts were asked to rate whether the intervention should be conducted exclusively in the ED, or alternatively, if could be conducted in a sub-acute healthcare centre. If an expert indicated an intervention suitable for a sub-acute centre, they were asked if it could be conducted in a: (1) urgent care centre, (2) walk-in medical centres, and (3) nurse practitioner led-clinics (multiple selections were permitted). These sub-acute centres were selected as they represented the most feasible centres patients could be redirected to when transported by paramedic services, their services target non-emergent events, are abundant in Ontario, and at present do not receive patients by ambulance. Standardized definitions of each destination were provided to minimize any heterogeneity in expert interpretation of a healthcare centres function. Additionally, descriptions of each healthcare centres staffing, imaging and non-clinical specialty service abilities were provided to increase inter-member consistency. Experts were provided a free-text comment space for each intervention to support their decision.

We determined consensus as any intervention receiving 70% or greater agreement amongst all experts for an individual health care centre (either ED or sub-acute centre). We collected all expert ratings from Round 1, extracted the data of individual reports and composed a general feedback form that contained aggregate percent agreement of all interventions reaching consensus, and those that did not. We hosted a videoconference debrief with the Delphi expert committee to share the results of Round 1, and to facilitate a discussion on the interventions that did not reach consensus.(19) We examined the comments of Round 1 for justifications of expert decisions, and to amend any framing of interventions that may have been unclear in Round 1 for Round 2. Experts were able to login to their questionnaire with their name, change their answers, and could not submit a second questionnaire to avoid duplication.

Round 2 of the modified Delphi included all ED interventions that did not receive consensus in Round 1. Expert ratings of Round 2 would serve as the final consensus level on the residual interventions.

## RESULTS

We identified 150 of the most frequently recorded Canadian Classification for Health Interventions conducted on non-emergent patients transported by paramedic services to an ED using the NACRS database. These interventions represented 95.5% (1 239 998/1 319 388) of all interventions recorded in this NACRS cohort of Ontario ED patients in the study period. All 150 interventions were included in Round 1 of the RAND/UCLA modified Delphi exercise, the majority of interventions belonging to CCI sections 1, 2 and 3 (137 / 150; 91.3%).

A total of 25 physicians who met the selection criteria were invited to participate by email, 21 accepted and consented to participate. Collectively 20 experts completed Round 1 and constituted the Delphi expert committee for this study, and completed 100% of each questionnaire. Experts were recruited from October 13 to November 5, 2020, and the modified Delphi consensus rounds occurred between November 6 and December 19, 2020. Figure 1 shows the flow of recruitment and modified Delphi Rounds in the study.

**Figure 1:**
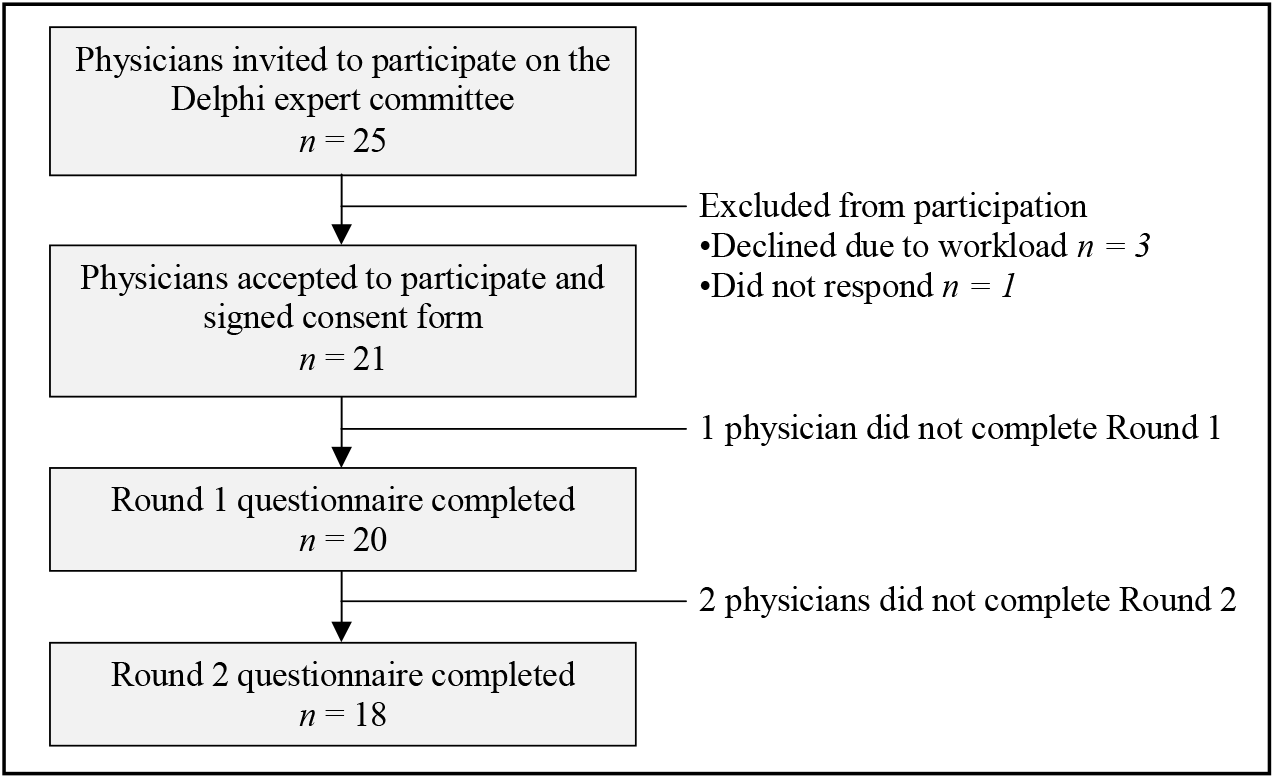
Study course of recruitment and 2 rounds of the RAND/UCLA modified Delphi consensus exercise.

The majority of experts were male (70%) and reported their primary medical practice as emergency medicine (80%), with the remaining as family medicine (15%) or both (5%). The characteristics of the Delphi expert committee are shown in Table 1.

**Table 1:** Demographic characteristics of the Delphi expert committee in the RAND/UCLA modified Delphi consensus exercise.

| **Characteristic** | **Experts, n=20**<br>**(n, %)** |
| --- | --- |
| **Gender** |  |
| Male | 14 (70%) |
| Female | 6 (30%) |
| **Province of practice (present)** |  |
| Ontario | 18 (90%) |
| Quebec | 2 (10%) |
| **Primary medical practice** |  |
| Emergency medicine | 16 (80%) |
| Family medicine | 3 (15%) |
| Both | 1 (5%) |
| **Length of practice, yrs** |  |
| 0 to 4 | 2 (10%) |
| 5 to 9 | 5 (25%) |
| 10 to 14 | 2 (10%) |
| 15 to 19 | 2 (10%) |
| 20 to 24 | 2 (10%) |
| 25 to 29 | 2 (10%) |
| 30 or greater | 5 (25%) |
| **Medical director, Ontario**<br>**paramedic practices** | 5 (25%) |

In Round 1, 139 interventions achieved a 70% consensus agreement amongst all experts (92.7%) for use in at least a single sub-acute healthcare centre. All interventions included in Round 1 were considered and rated by all 20 participating experts. The remaining 11 interventions which did not achieve consensus were included in Round 2, of which all were CCI section 1 interventions (i.e., physical/physiological therapeutic interventions). In Round 2, seven additional interventions reached consensus from experts for use in at least a single sub-acute healthcare centre, with the remaining four not reaching consensus in this study. Two of the experts that complete Round 1 did not complete round 2. Consensus results in the RAND/UCLA modified Delphi exercise are shown in summary by round and healthcare centre in Table 2.

**Table 2:**
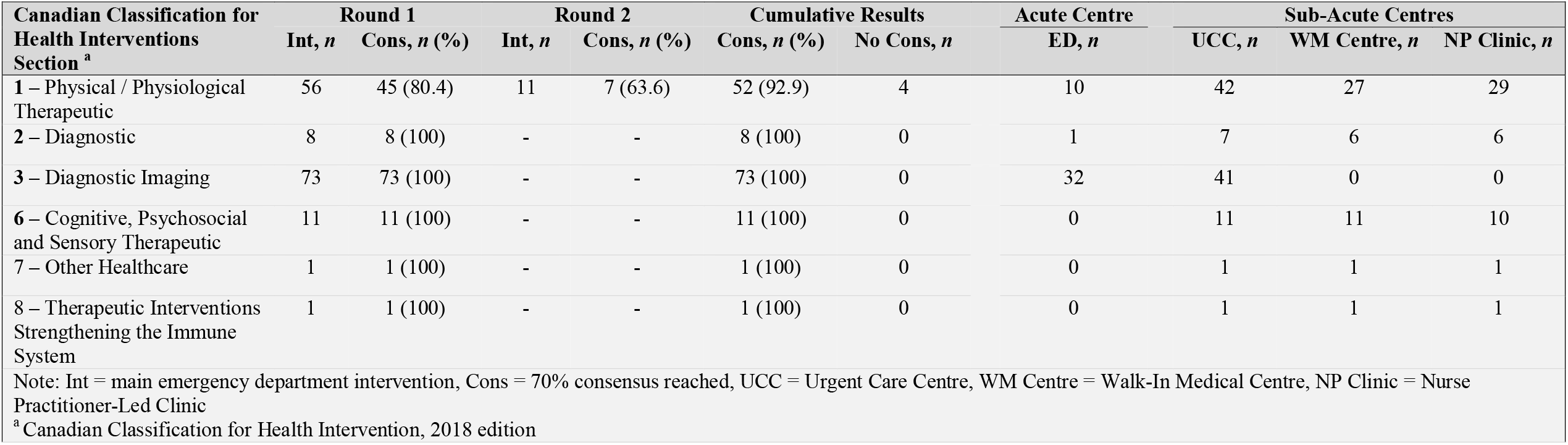
Emergency department interventions receiving consensus through each round of the RAND/UCLA modified Delphi, and final results of healthcare centre that could conduct the intervention.

Of ED interventions that achieved overall consensus, 103 (68.7%) were rated from experts for their suitability for a sub-acute healthcare centre. Of the 47 remaining, 43 interventions (28.7%) were rated as only appropriate for the ED, and four interventions did not reach consensus (2.6%). All 103 intervention were deemed suitable for an urgent care centre, of which 46 interventions were suitable for a walk-in medical centre, and 47 for a nurse practitioner-led clinic. Of interventions requiring diagnostic imaging (Section 3), all magnetic resonance imaging (MRI) or computed tomography (CT) imaging were identified as only suitable for the ED, while the remaining two imaging categories (x-ray, ultrasound) were all rated appropriate for urgent care centres. All interventions of CCI Sections 6, 7 and 8 were determined to be appropriate for sub-acute healthcare centres, and nearly all inventions of Section 2. The four interventions that did not receive consensus ranged in rating of 50-66% amongst expert agreement. All interventions receiving consensus for any of the three sub-acute healthcare centres are shown in Table 3, and results of the RAND/UCLA modified Delphi agreement ratings for all interventions are shown in Table 4 (Appendix).

**Table 3:**
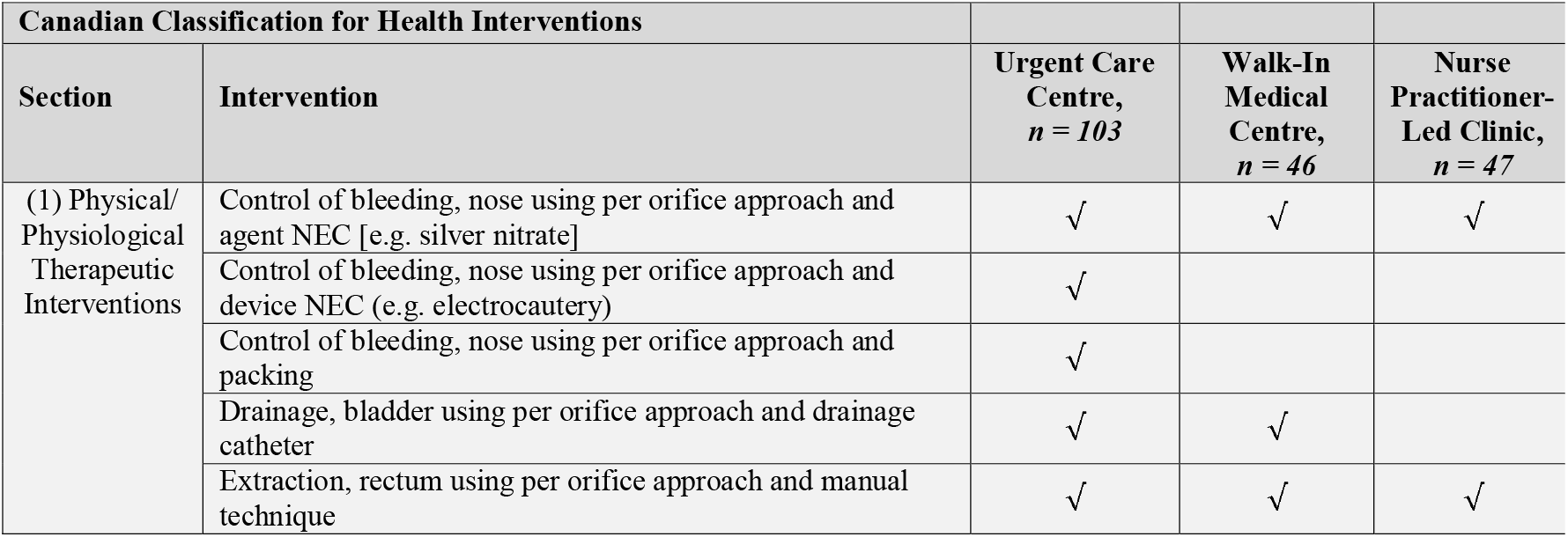
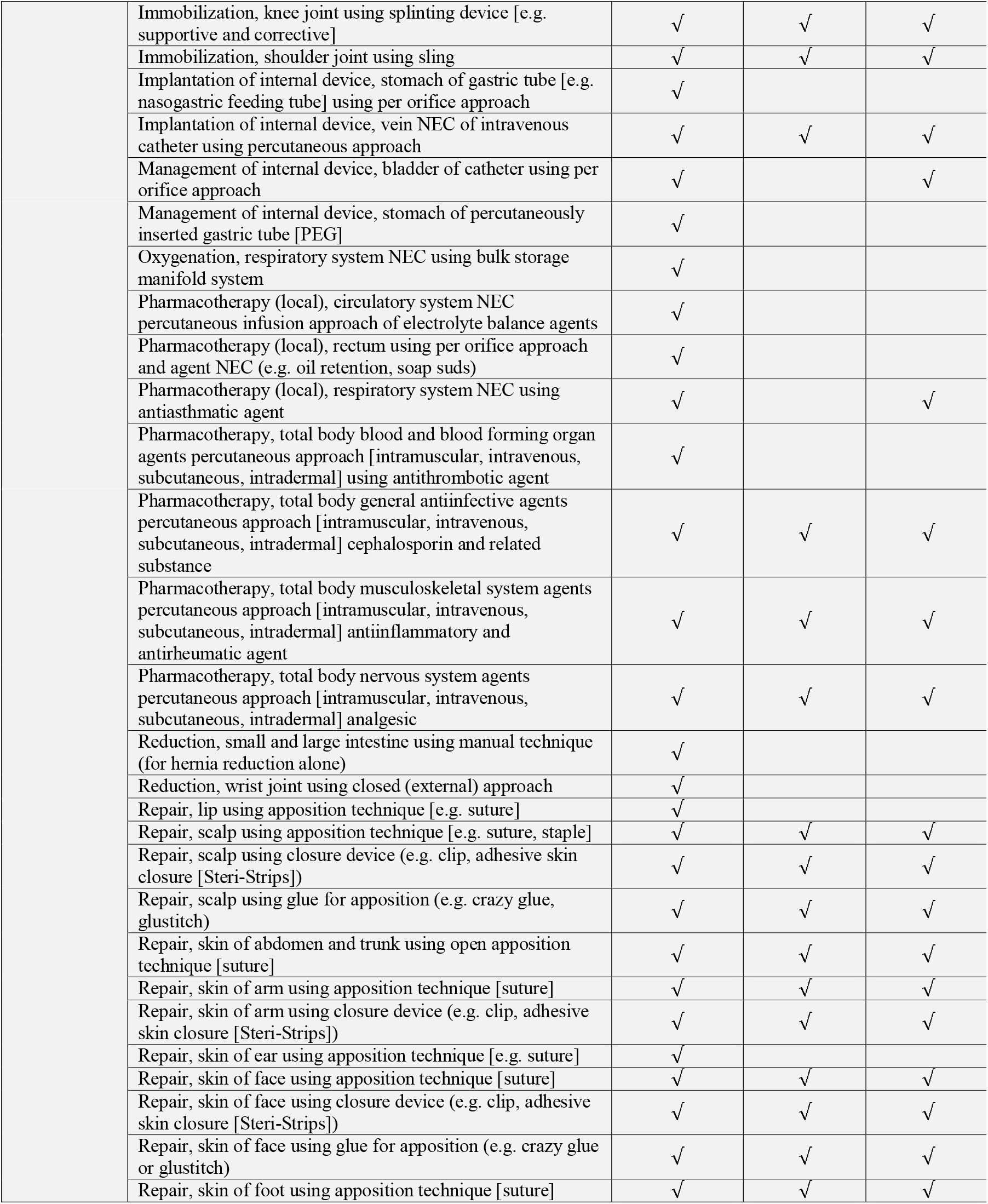
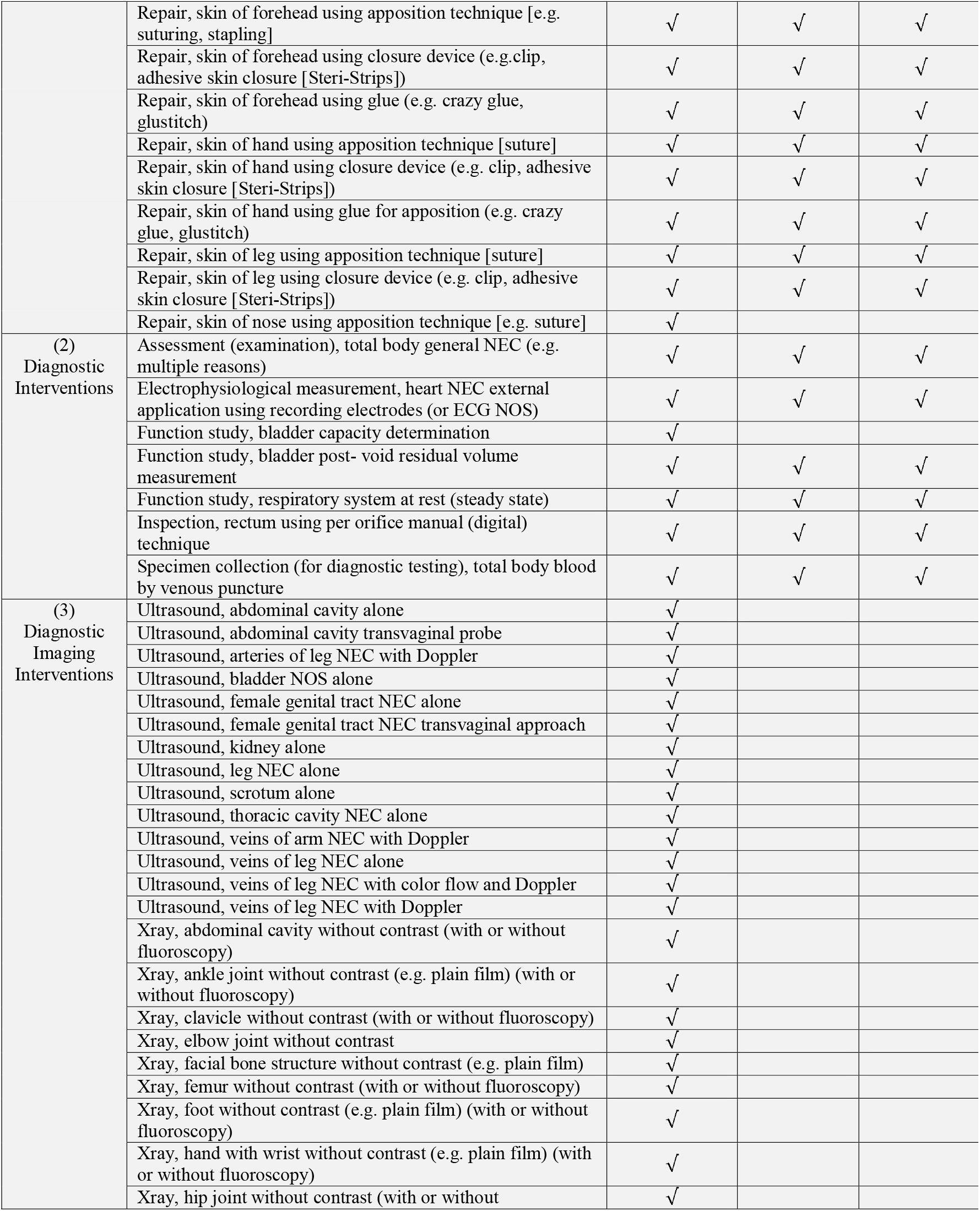
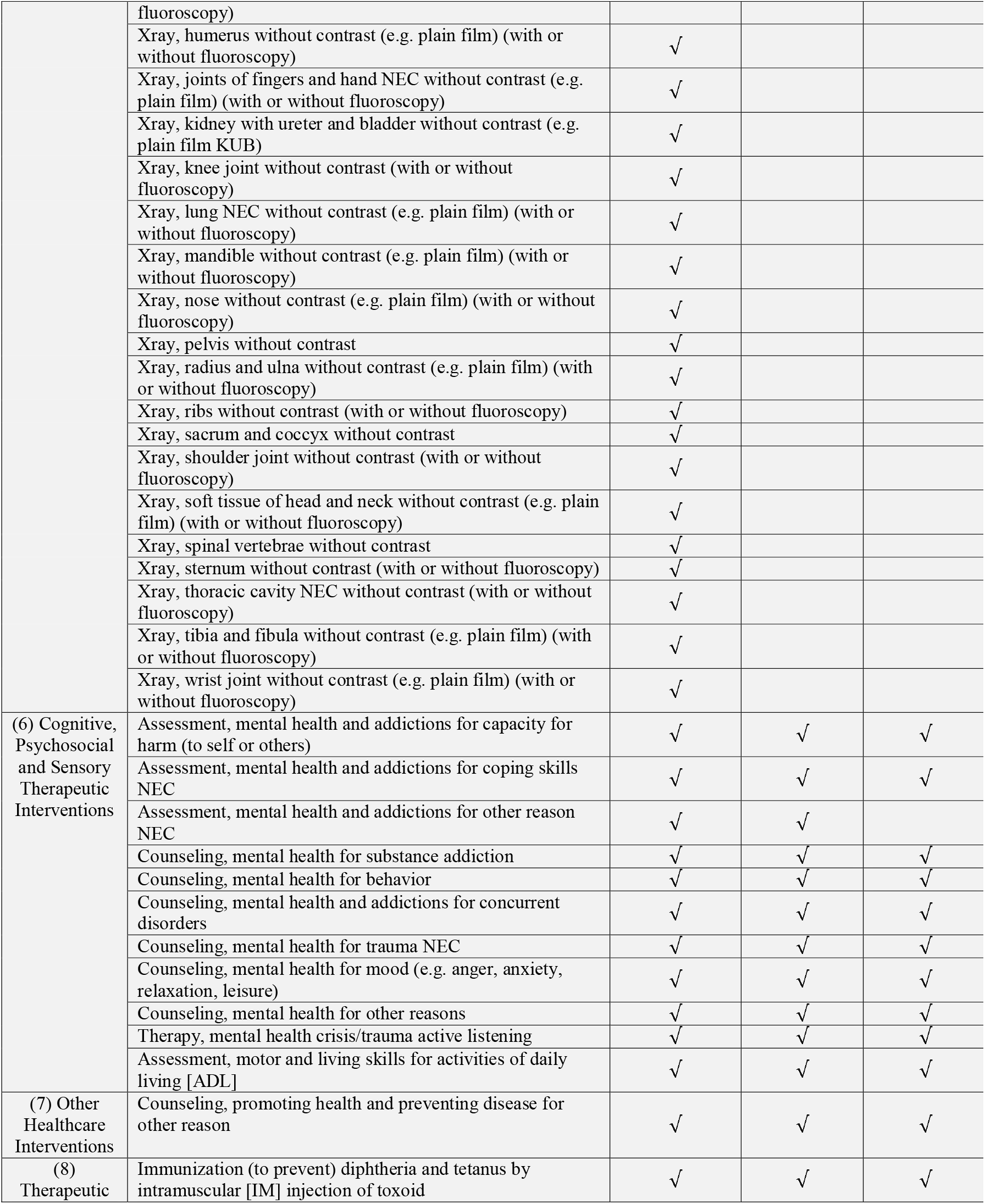
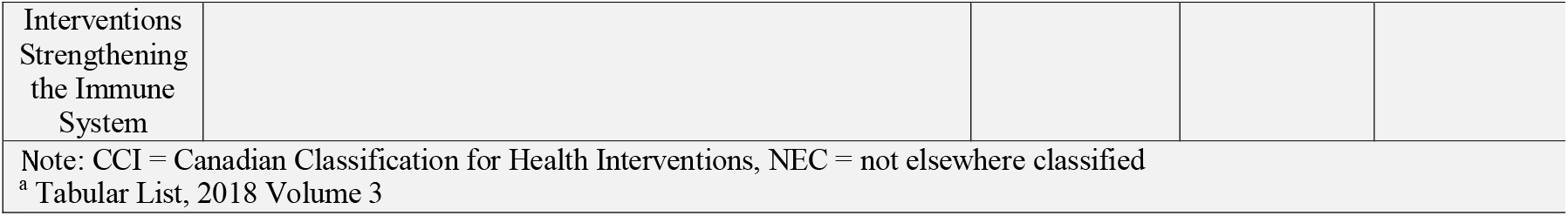
Emergency department interventions on non-emergent paramedic transported patients deemed suitable for sub-acute healthcare centres, shown by care centre.

Expert comments highlighted that their selections for healthcare centres were based the underlying assumption that the clinicians of the centres had adequate training and experience with a given intervention. Experts commented that interventions rated for ED suitability only, were largely evaluated that way based on concerns for patient safety if the procedure was unsuccessful. Interventions that did not receive consensus in Round 2 were identified as requiring an element of sedation and a second physician for safety and competence purposes, although this was not agreed upon by all. Some experts emphasized their ratings do not infer any direction for clinical guidance based solely on this study, as an intervention alone does not provide enough specific information to inform care planning without further contextualization.

Experts rated the majority of all interventions as suitable for urgent care centres. Their justification included the urgent care centre’s ability to be equipped with the resources required to conduct the interventions (e.g., diagnostic equipment, technicians trained to use the equipment). Comments suggested that walk-in medical centres and nurse practitioner-led clinics may not have access to the same diagnostic imaging recourses, leading experts to deter from selecting these centres as appropriate. Experts did not cite practitioner skill as a limiting factor in rating decisions; however, they did acknowledge some interventions are based on physician or practitioner preference. Some experts recommended there could be other centres that are more appropriate for psychological interventions (e.g., facilities that focus solely on mental health), but were not included in the study. The videoconference held between rounds 1 and 2 contributed insights into nurse practitioner-led clinics receiving more consensus on interventions than walk-in medical centres. Experts were also unsure if walk-in centres were staffed with a single clinician, whereas for nurse-practitioner clinics they felt were typically staffed with more than one, which led to an increased comfort with rating interventions that are more time-consuming or required extended monitoring.

## INTERPRETATION

There is consensus by primary and emergency care physicians on what clinical interventions commonly performed in the ED for non-emergent paramedic transported patients are suitable for alternative sub-acute healthcare settings. Specifically, 68.7% of included ED interventions were rated as suitable for conduction in urgent care centres, 30.7% in walk-in medical centres and 31.3% for nurse-practitioner led clinics, while 2.6% did not receive consensus.

Our results are consistent with previous literature that suggests urgent care centres and similar sub-acute centres can be reasonable avenues for treatment of non-emergent patient conditions who would otherwise be directed to the ED.(11,24) There is an absence of evidence that measures the appropriateness of which ED interventions could be conducted in sub-acute settings, as most articles analyze patient conditions, diagnostics and medication administration.(11,12,25) Previous literature describes 13.7% to 27.1% of all ED patients could be safely managed by urgent care, however do not report which interventions were conducted.(11) Ample literature describes the use of sub-acute centres to offset ED use, however focus heavily on outcomes of patient satisfaction and cost avoidances(11,24), when quality of care, care received and simulation modeling may be more important indicators for supporting paramedics redirection models.(26,27)

The majority of all included interventions found to be appropriate for sub-acute centres acknowledges the confidence that study experts have in a sub-acute centres ability to conduct emergency department interventions. Of interventions that were rated for ED only, many required sedation practices, intensive monitoring, or advanced emergency physician skills. The four interventions that did not receive consensus all shared the same intervention procedure of using a reduction technique to treat an injury. Of Section 3 interventions involving diagnostic imaging, equipment was determined as the limiting factor (not injury site or physician interpretation).

An overarching goal of our study was to determine if consensus on which ED interventions could be performed elsewhere, such that an epidemiological patient classification could be constructed to inform redirection by paramedics. We recognize that interventions alone are insufficient considerations for such redirection programs. However, in combination with other indicators (e.g., contextualized patient features) and supports (e.g., education), knowledge of interventions suitable for sub-acute healthcare centers will support the construction of a patient classification model for paramedic-initiated redirection from the ED. Future research is required to incorporate additional patient and administrative information into a classification in order to provide contextualization before evaluating its validity for clinical guidance. The results of this study contribute evidence towards informing the circumstances (in part) in which paramedic service-based programs intended to support redirecting ED bound patients may be feasible and appropriate.

## LIMITATIONS

An inherent limitation of using secondary administrative datasets is the completeness of the procedural fields. Our dataset included 63.7% completeness of the main interventions field in NACRS (1 319 388 / 2 070 260), to which this is expected as patients admitted to hospital may have their ED interventions recorded in the Discharge Abstract Database as opposed to NACRS. In other instances, there was no intervention completed during the visit, or the intervention was not recorded. Our cohort size remained large and is trustworthy based on our study objectives.

Individual judgements may be subjective given an expert’s own evaluation with safety in selecting healthcare centres. This limitation was minimized in the study design to include only physicians with adequate knowledge of emergency and primary care practices in Ontario, the Delphi committee contained a high number of experts, and a detailed description of each healthcare centre was provided. The findings of this study may not be generalizable in settings where payment structures for healthcare, accessibility to sub-acute care or ambulance availability are different. Additionally, our research was specific in terms of population (adult, non-emergent, ambulance) and only included ED interventions, without taking into consideration additional clinical details.

## CONCLUSION

With a continued increase in the proportion of non-emergent or low acuity patients transported to EDs by paramedic services, it is important to explore features supporting redirection programs such that their impact on patient and ED utilization outcomes can be examined. A majority of ED interventions conducted by physicians on non-emergent patients transported by paramedic services were identified as suitable for conduction in sub-acute healthcare centres including urgent care centres, in walk-in medical centres and nurse practitioner-led clinics. While focusing on interventions alone has limitations, these results suggest there may be a patient population that may be suitable for redirection programs by paramedic services in Ontario as a way of countering the ED crisis. These results may contribute evidence to inform construction of a patient classification system for non-emergent patients for use by paramedic services that (a) could potentially be used to prevent ED visits and (b) better align paramedic services with patient needs. Future research is required to augment our findings with additional patient and hospital contextualization toward such a classification system.

## Data Availability

All data are included in the manuscript and appendix.

## Completing Interests

None declared.

## Acknowledgements

The authors would like to thank all 20 physicians of the Delphi expert committee who participated in the RAND/UCLA modified Delphi exercise and for their valuable contributions.

## Contributions

RPS and APC led the conceptualization of the study methodology. RPS designed the study, conducted the RAND/UCLA modified Delphi exercise, and drafted and revised the manuscript. WT, AW and LEG made contributions to the design of the study, methods, interpretation and manuscript. All authors critically revised the manuscript, approved the final version and agreed to act as guarantors.

## Funding

The investigators received no specific funding for this study.

## Ethics

This study received a research ethics board exemption waiver from the Hamilton Integrated Research Ethics Board; review reference 2020-11451-GRA.

## APPENDIX

**Table 4:**
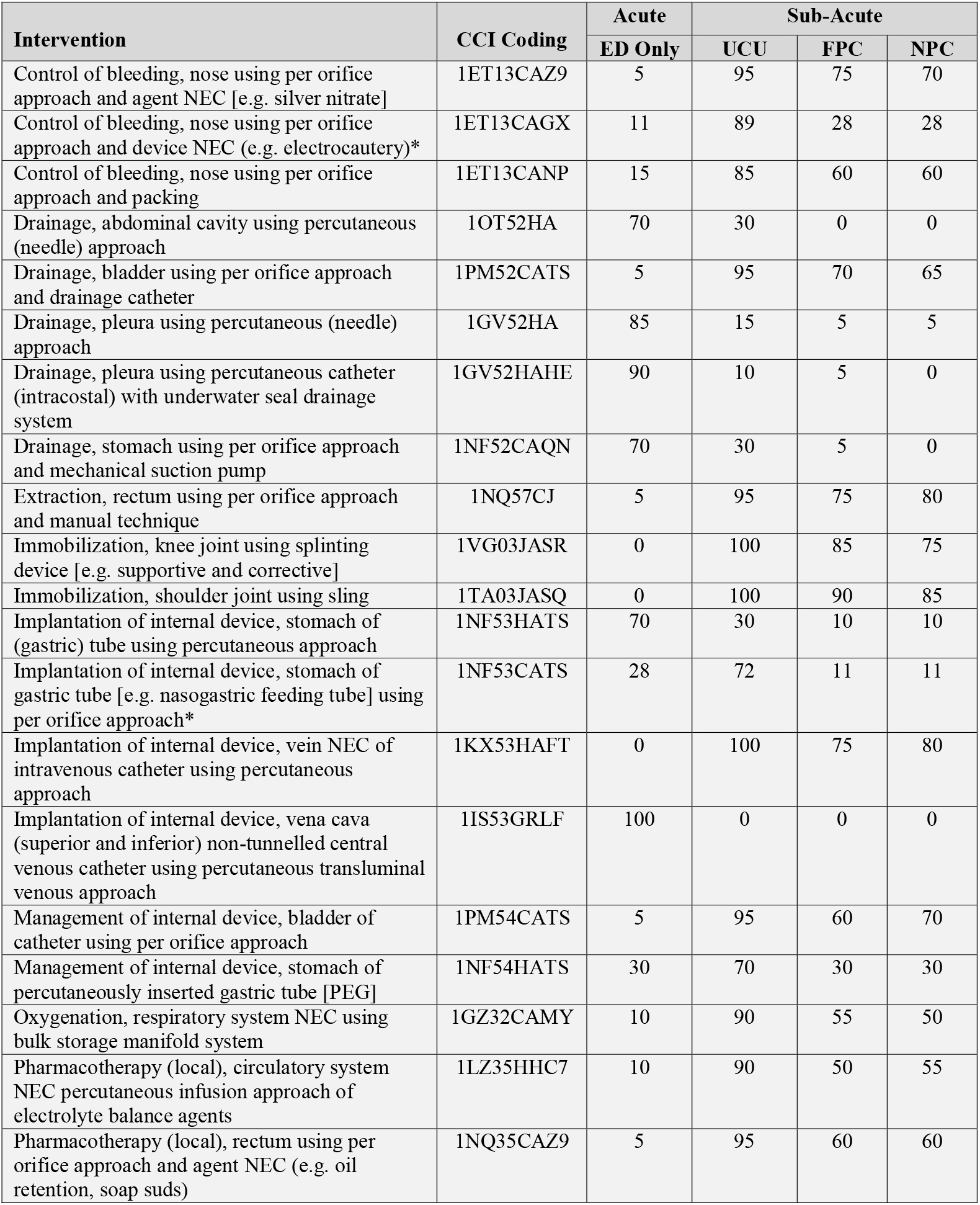
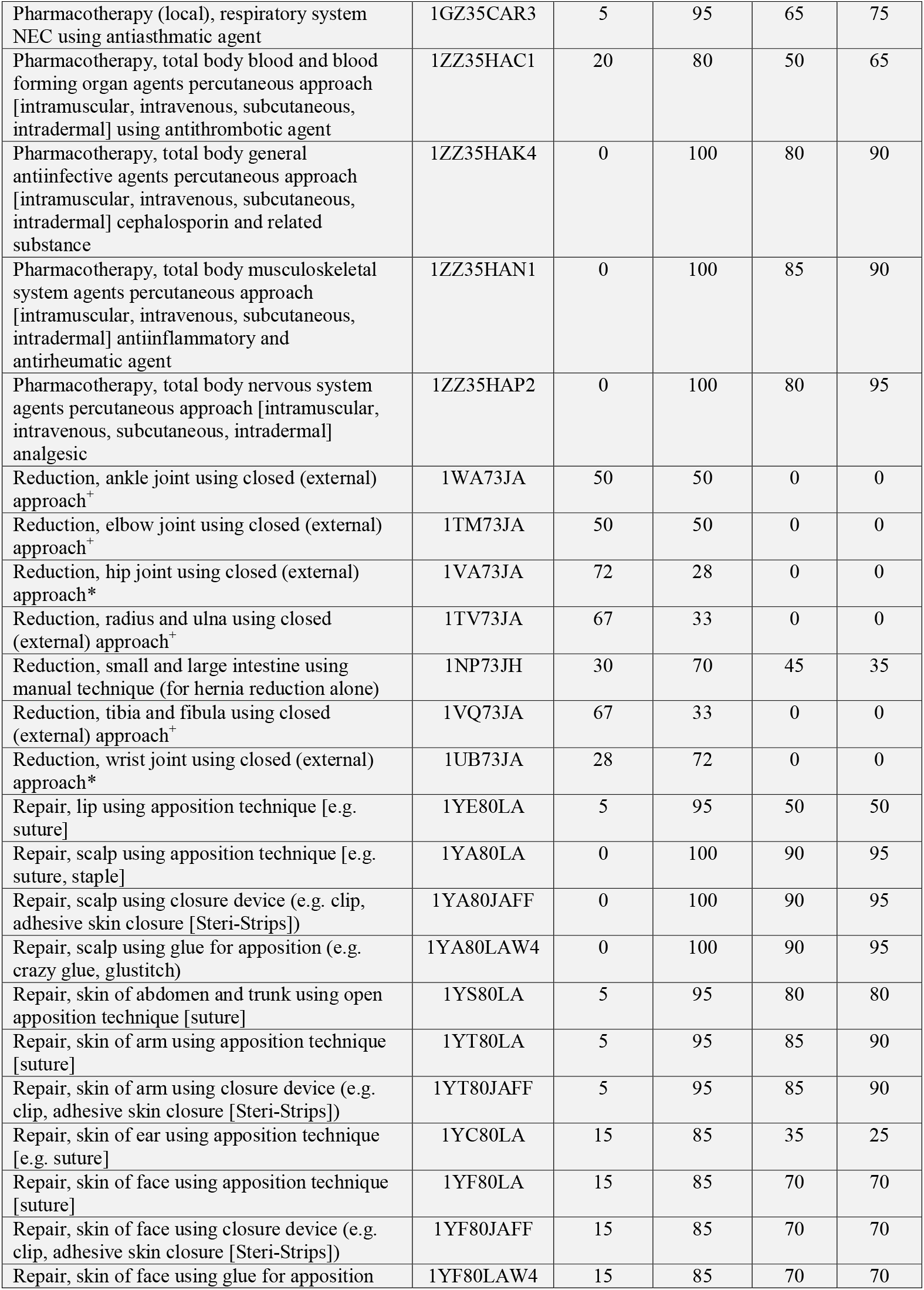
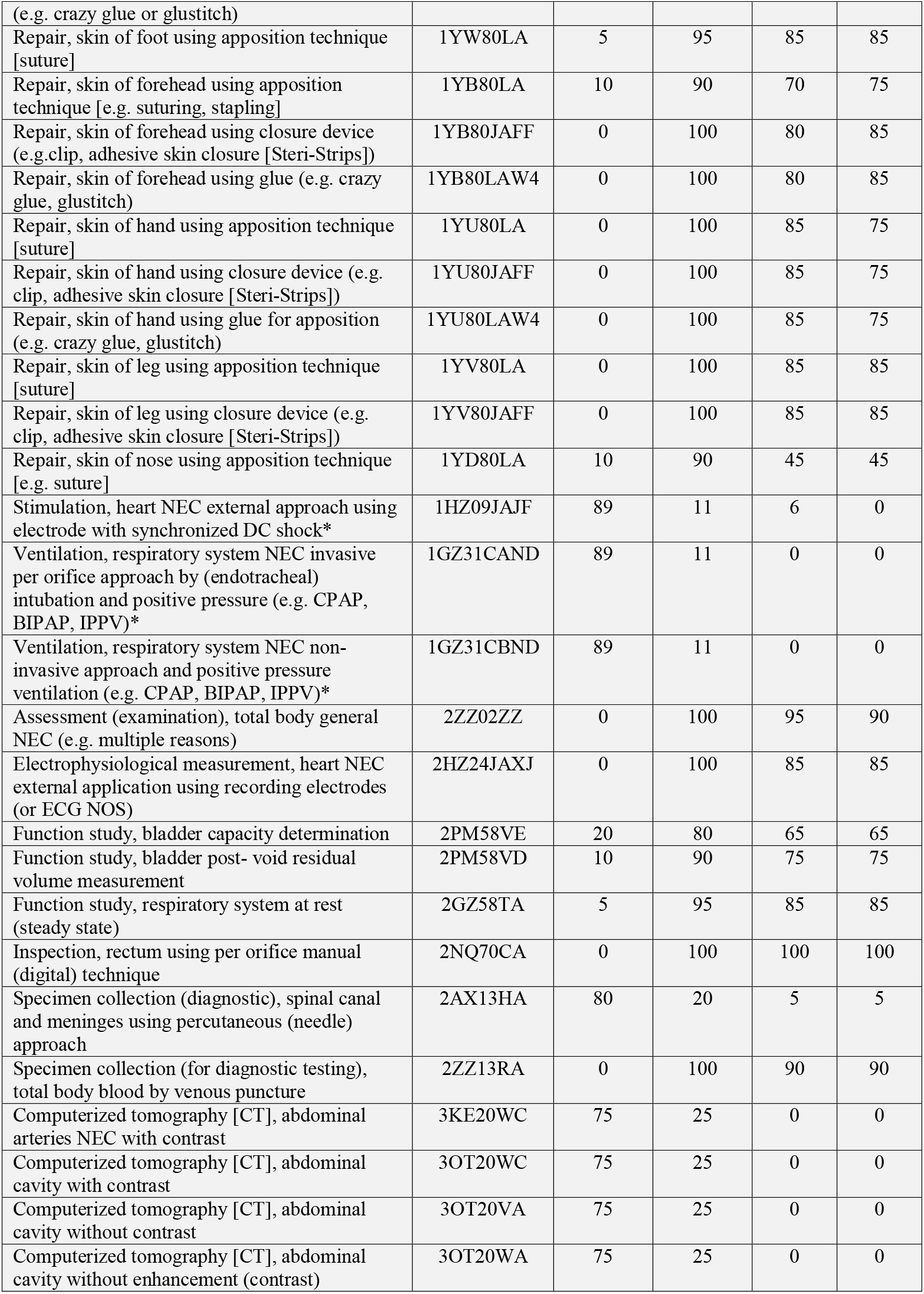
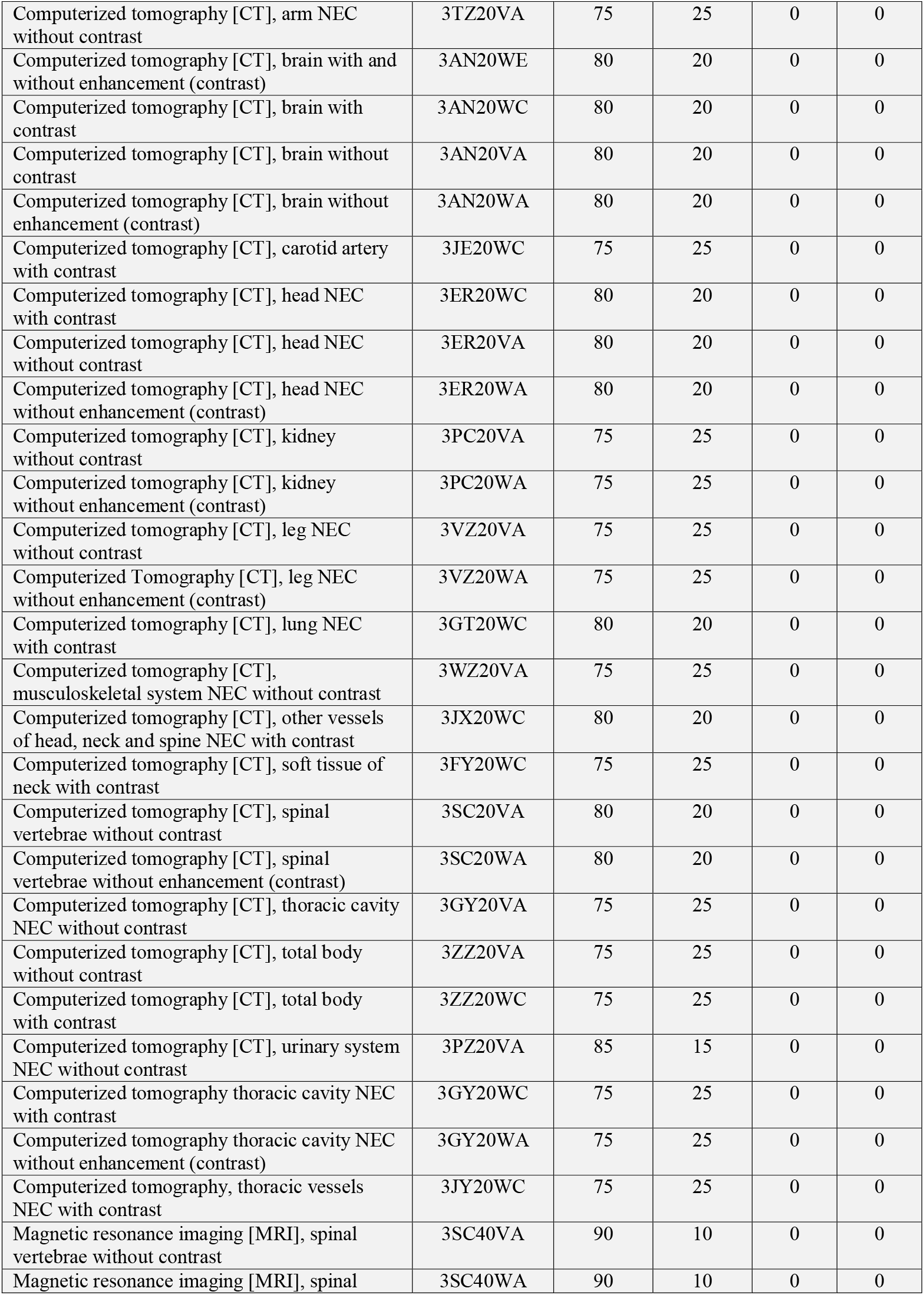
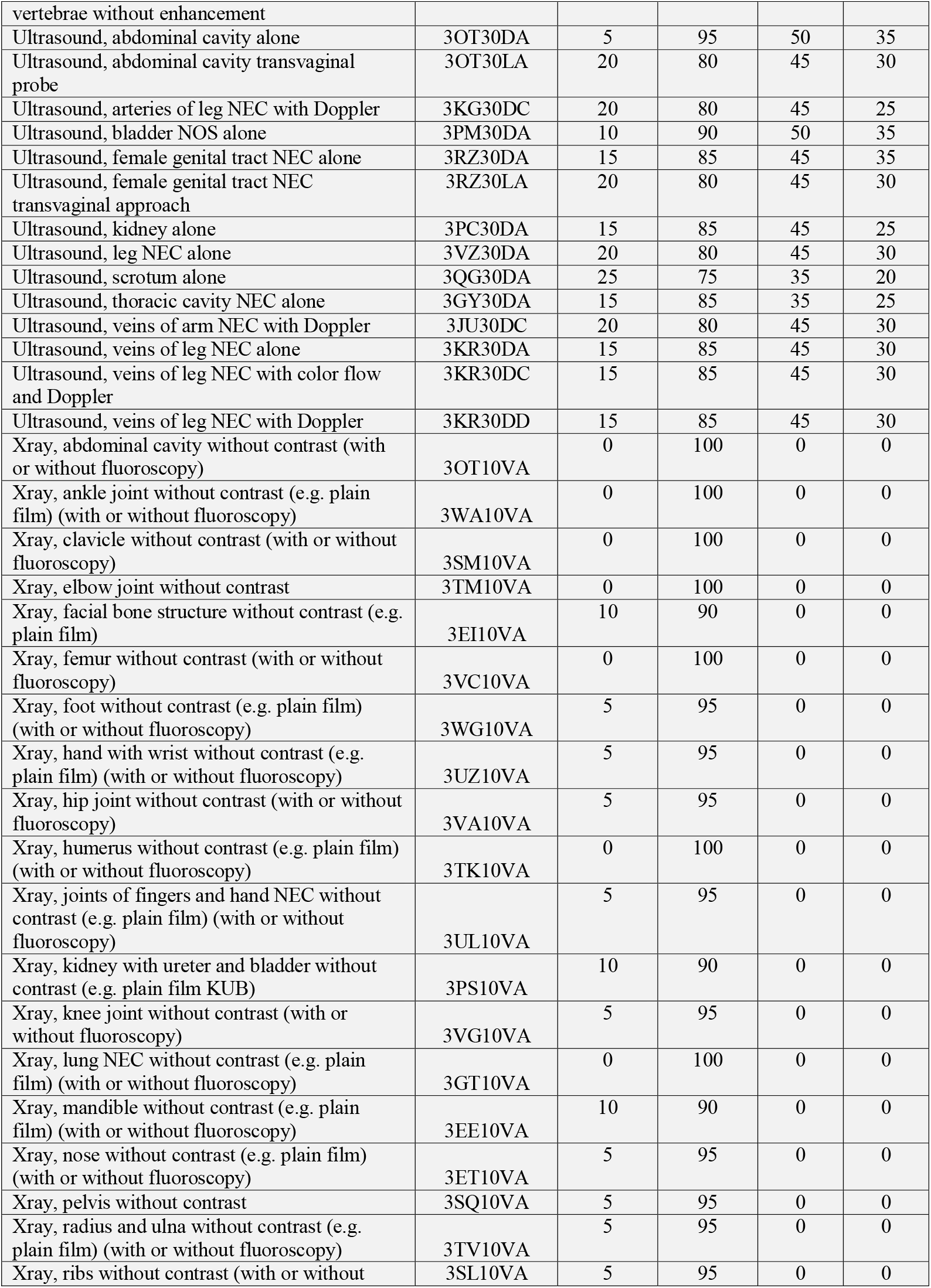
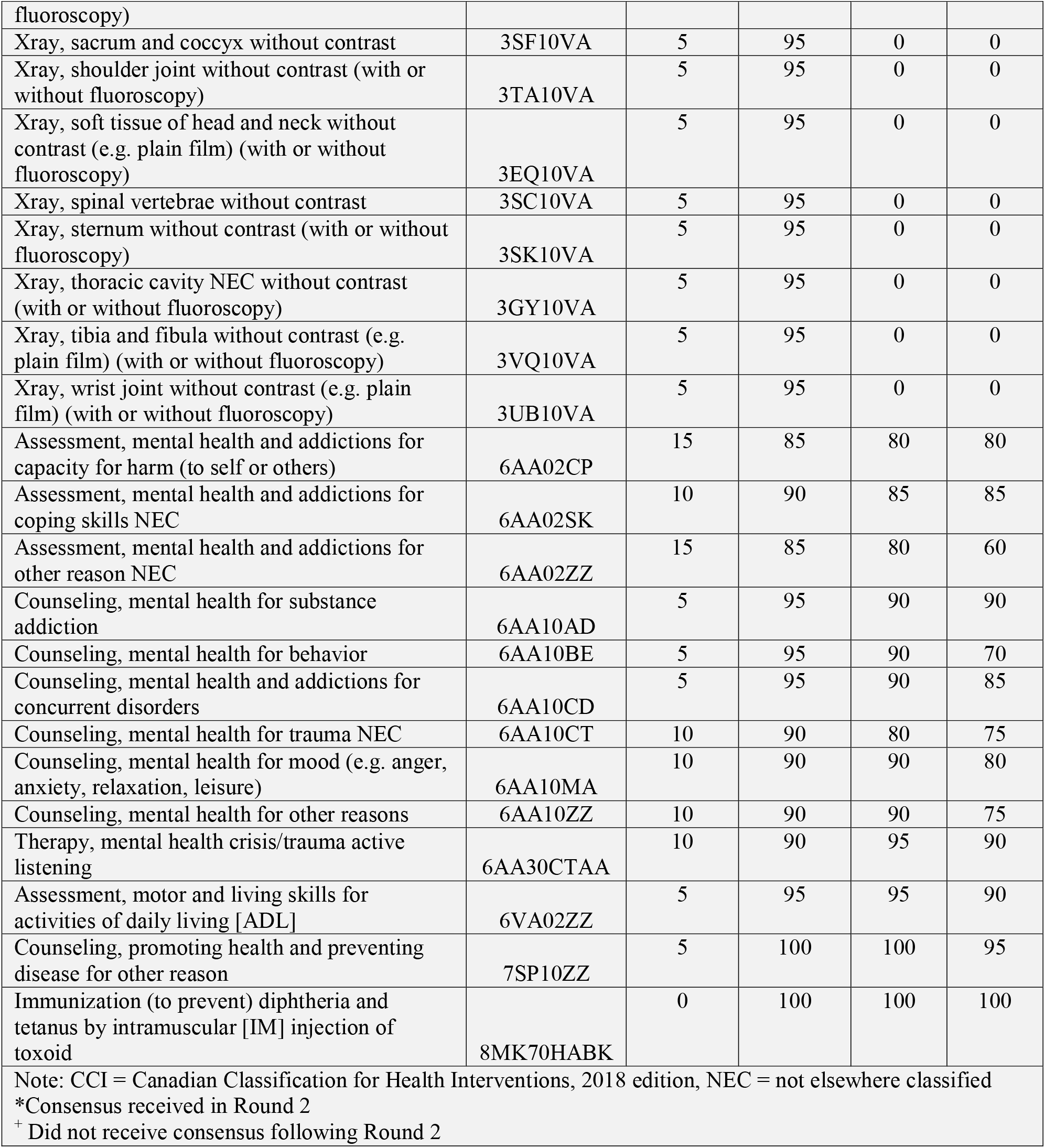
Intervention consensus of the Delphi expert committee from the modified Delphi exercise, shown as percentages.

## Notes

### Competing Interest Statement

The authors have declared no competing interest.

### Clinical Trial

ISRCTN22901977.

### Clinical Protocols

https://bmjopen.bmj.com/content/11/1/e045351

